# Screening plans for SARS-CoV-2 based on sampling and rotation: an example in the school setting

**DOI:** 10.1101/2021.02.10.21251502

**Authors:** Michela Baccini, Giulia Cereda

**Affiliations:** Department of Statistics, Computer Science, Applications (DISIA), University of Florence, Florence, Italy

## Abstract

Screening plans for prevention and containment of SARS-CoV-2 infection should take into account the epidemic context, the fact that undetected infected individuals may transmit the disease, and that the infection spreads through outbreaks, creating clusters in the population. In this paper, we compare the performance of six screening plans based on poorly sensitive individual tests, in detecting infection outbreaks at the level of single classes in a school context. The performance evaluation is done by simulating different epidemic dynamics within the class during the five weeks following the day of the first infection. The plans have different costs in terms of number of individual tests required for the screening and are based on recurrent evaluations on all students or subgroups of students in rotation. Especially in scenarios where the rate of contagion is high, at an equal cost, testing half of the class in rotation every week appears to be better in terms of sensitivity than testing all students every two weeks. Similarly, testing one-fourth of the students every week is comparable with testing all students every two weeks, despite the first one is a much cheaper strategy. In the presence of natural clusters in the population, testing subgroups of individuals belonging to the same cluster in rotation may have a better performance than testing all the individuals less frequently. The proposed simulations approach can be extended to evaluate more complex screening plans than those presented in the paper.

## INTRODUCTION

Since the beginning of the COVID-19 emergency in the early 2020, the importance of implementing extensive screening procedures to prevent or slow down the spread of the SARS-CoV-2 infection has been emphasized [1, 2] and, in light of the threat of new variants of the virus that could be more widespread and the critical issues related to the rapid implementation of vaccination plans [3, 4], it still seems early to consider extensive surveillance strategies on the population (or specific subgroups of it) no longer necessary. Pharmaceutical industries have produced tests of various nature and cost, which have been proposed and used in screening plans aimed at early detection of asymptomatic or paucisymptomatic individuals in specific populations. The ability of these tests to correctly classify the single patient as infected or not has been widely discussed and debated, sometimes overshadowing the necessity that screening plans account for strengths and limitations of the used tests and are tailored to the specific context in which they are applied [5, 6].

When dealing with a screening plan for an infectious disease, two points should not be overlooked:

- undetected infected subjects can transmit the disease;
- the infection usually spreads in small outbreaks, creating clusters in the population (families, classes, work colleagues).

For these two reasons, screening plans similar to those implemented in the case of non-communicable diseases could lead to suboptimal results in terms of cost-benefit ratio. Furthermore, it is crucial that the screening procedures are assessed accounting for the actual epidemic context, including the strength of contagion [7].

In this paper, we compare through simulations alternative strategies designed for screening in schools, based on repeated tests to be performed at regular time intervals on all students, or on tests to be performed in rotation on subgroups. The comparison, which refers to a single class of *N* = 24 students - the class is here defined as a group of students who attend the same course each day at university or school - takes into account the epidemic context and is performed under alternative scenarios of epidemic spread.

Although the simulation analysis refers to school settings, the methods adopted and the results obtained are valid for any context in which there are natural clusters of individuals, within which contagion could spread starting from a single initial infection.

## METHODS

Our objective is to early identify infections in school settings, in order to quarantine the classes where there is at least one infected student. Let us suppose that individual tests are performed on all students in the class or on a subset of them. The class is considered positive if at least one of the individual tests is positive; it is considered negative if no individual tests are positive. Therefore, at the class level, the sensitivity of the test is defined as the probability that the class is positive given that there is at least one infected student present. Specificity is the probability that the class is negative given that there are no infected students in it.

In our analysis, we assume that the individual tests used in the screening procedure have maximum specificity and sensitivity equal to *p*. The assumption of maximum specificity of the individual test implies maximum specificity at the class level and rules out those situations where false positives may lead to quarantining classes when not necessary. This assumption allows us to simplify subsequent calculations without compromising evaluations regarding the ability of the proposed plans to detect outbreaks.

### Screening plans

We consider six screening plans (Figure 1), which differ from each other for the time interval between consecutive evaluations on the class and number of students involved in each of them:

**Figure 1.**
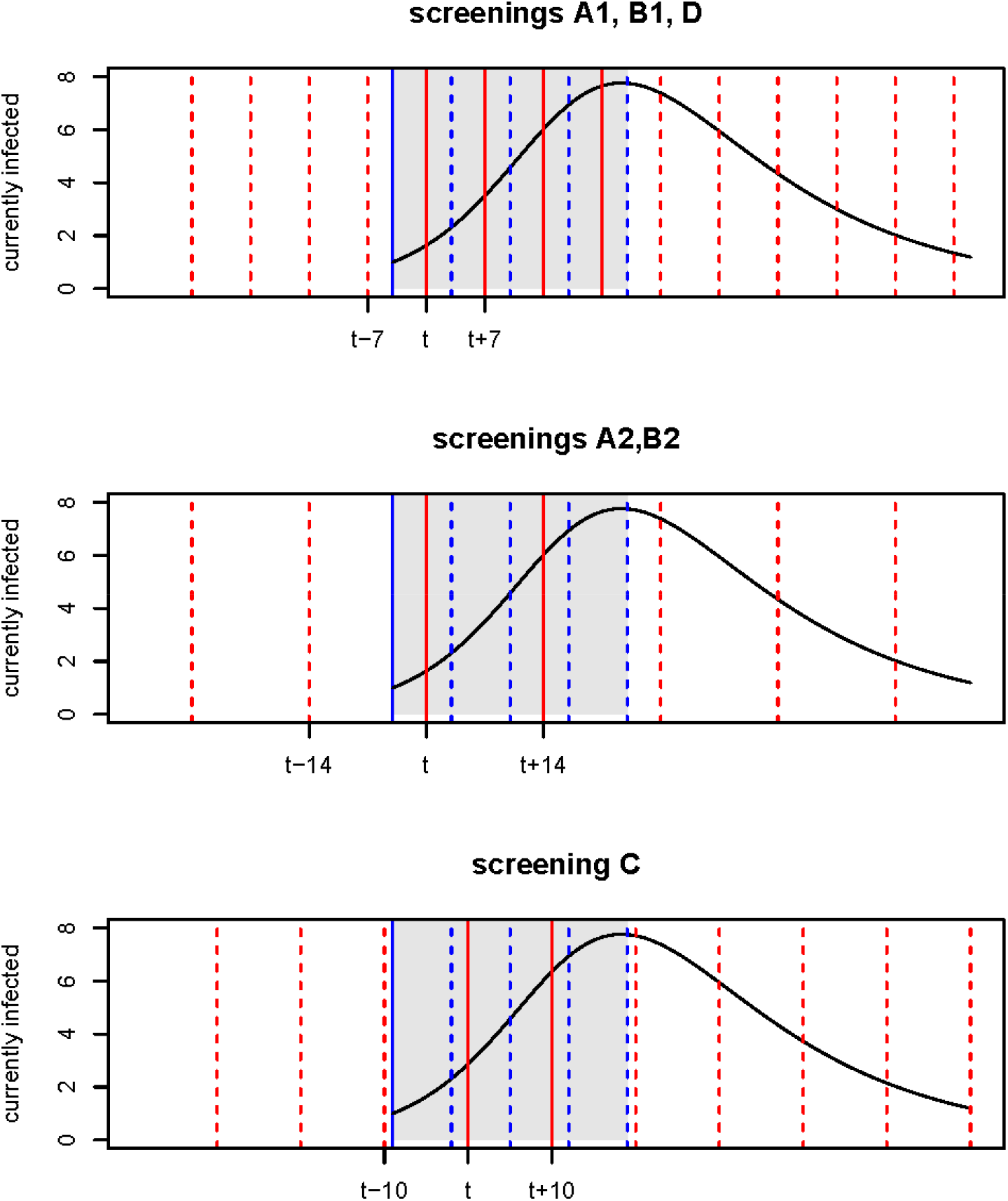
Example of an epidemic curve and its intersection with the screening plans (A1, A2, B1, B2, C, D). Dashed and solid red lines: evaluations carried out on the class according to the screening plans; Solid blue line: beginning of the epidemic; Grey area: time window on which the assessment is made (4 weeks from the beginning of the epidemic); Dashed blue lines: 1, 2, 3 and 4 weeks from the beginning of the epidemic; Solid red lines: evaluations carried out on the class within four weeks from the beginning of the epidemic.

- Plan A1. Individual tests on all students of the class every week;
- Plan A2. Individual tests on all students of the class every 2 weeks;
- Plan B1. Individual tests every week on 1/2 of the students of the class, in rotation;
- Plan B2. Individual tests every 2 weeks on half of the students of the class, in rotation;
- Plan C. Individual tests every 10 days on 1/3 of the students of the class, in rotation;
- Plan D. Individual tests every week on 1/4 of the students of the class, in rotation.

Plan A1 guarantees the best performance in terms of surveillance but requires more resources compared to the others. Plans B1, B2, C, and D have an additional element of risk compared to plan A2 because they test each time sub-samples of the students of the class. Plans B1, C and D allow more frequent monitoring of the class compared to plan A2 and B2.

We assume *p*=0.7, which is close to the average sensitivity reported by OECD for antigenic tests [6]. Then, in order to perform a sensitivity analysis, we assume *p*=0.9 as well.

### Epidemic scenarios

We assume that an infected student in the class may generate new infections among his/her classmates. Let us suppose that one of the *N* students is infected on day 1. The epidemic dynamic within the class can be simulated by using a compartmental model [8], where the contagion strength depends on the average time of infectivity *T* and on the basic reproduction number *R*_0_, which is the number of secondary infections generated from the first infected student in the class. In particular, we assume that, on average, each infected student may spread the contagion in the class for *T* days, still remaining detectable as infected for 4 weeks [9].

We consider different epidemic contexts, characterized by different combinations of *R*_0_ and *T* (*R*_0_=1.1, 1.5, 3, 5 and *T*=7, 14, 21, for a total of 12 scenarios) [10], and we apply on each of them the six screening plans. The ratio *β*=*R*_0_*/T* is proportional to the contagion rate; combinations of *R*_0_ and *T* which result in the same *β* generate the same epidemic dynamic, net of stochastic variability. This is the case for *R*_0_=2, *T*=14 and *R*_0_=3, *T*=21.

### Simulations

Separate simulations, for a minimum of 7000, are performed for each combination of *R*_0_ and *T* and each screening plan. We assume that the number of new infections and the number of infected that become not infectious at time *t* in the class, *I*_*new*_(*t*) and *R*_*new*_(*t*), follow Binomial distributions:

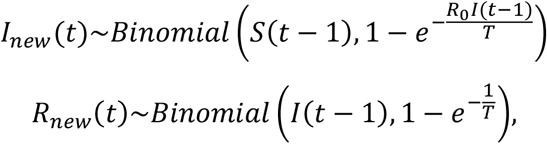

where *S*(*t* − 1) and *I*(*t* − 1) are the number of susceptible students and the number of infectious students at time *t* − 1, respectively. We further assume that the groups, when required by the screening plan, are randomly generated, that the probability of becoming infected for a susceptible subject does not depend on the group to which he/she belongs, and that the new infections at each time are randomly distributed among the groups. Let us suppose that *g*=2 groups of students *G*_1_ and *G*_2_ have been created and the susceptible individuals in the two groups at time *t* are *S*_1_(*t*), *S*_2_(*t*). The number of new infections in *G*_1_ is sampled from a Hypergeometric distribution:

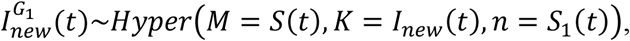

where *M* is the population size, *K* is the number of successes in the population, and *n* is the number of draws. Then the number of new infections in *G*_2_ is obtained as difference: 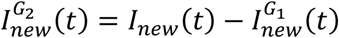. In general, if *g*>2 the number of new infections in the groups is obtained by sampling from a Multivariate Hypergeometric distribution.

The results of the individual tests on different individuals are assumed to be independent and follow a Bernoulli distribution with parameter *p*. According to the assumption of maximum specificity, the result of the test on the not infected students is always negative. In the simulations, we allow the epidemic to originate at any time between two consecutive assessments of the class and we focus on a time window of 4 weeks from the first infection (Figure 1) to evaluate the performance of the screening plan in terms of:

- probability of detecting the outbreak at 7, 14, 21 and 28 days since its onset;
- total number of infection-days which are left undetected by the screening plan in the 4 weeks time window. An infection-day is here defined as a day spent by a subject in the infectious status, thus a day in which he/she can spread the contagion.

## RESULTS

Figure 2 reports, for each scenario, the probabilities of a positive result on the class within 7, 14, 21 and 28 days from the beginning of the epidemic. The curve describes the overall performance of the screening plans in detecting the presence of infections in the class. As expected, plan A1 guarantees the best performance in terms of epidemic detection. The cumulative probabilities for plans A2 and B1 are very similar in scenarios where the infection spreads slowly within the class (upper left quadrant), while plan B1 seems to have better relative performance compared to A2 in high-epidemic spread contexts (bottom right quadrant). Screening plans B2, C and D have the worst performance if the infection spread is low, but they reach good results in high-risk scenarios. Plan D seems to detect infections within 2 weeks from the beginning of the epidemic with a probability between 70% and 80% in scenarios where *β* ≥ 0.36.

**Figure 2.**
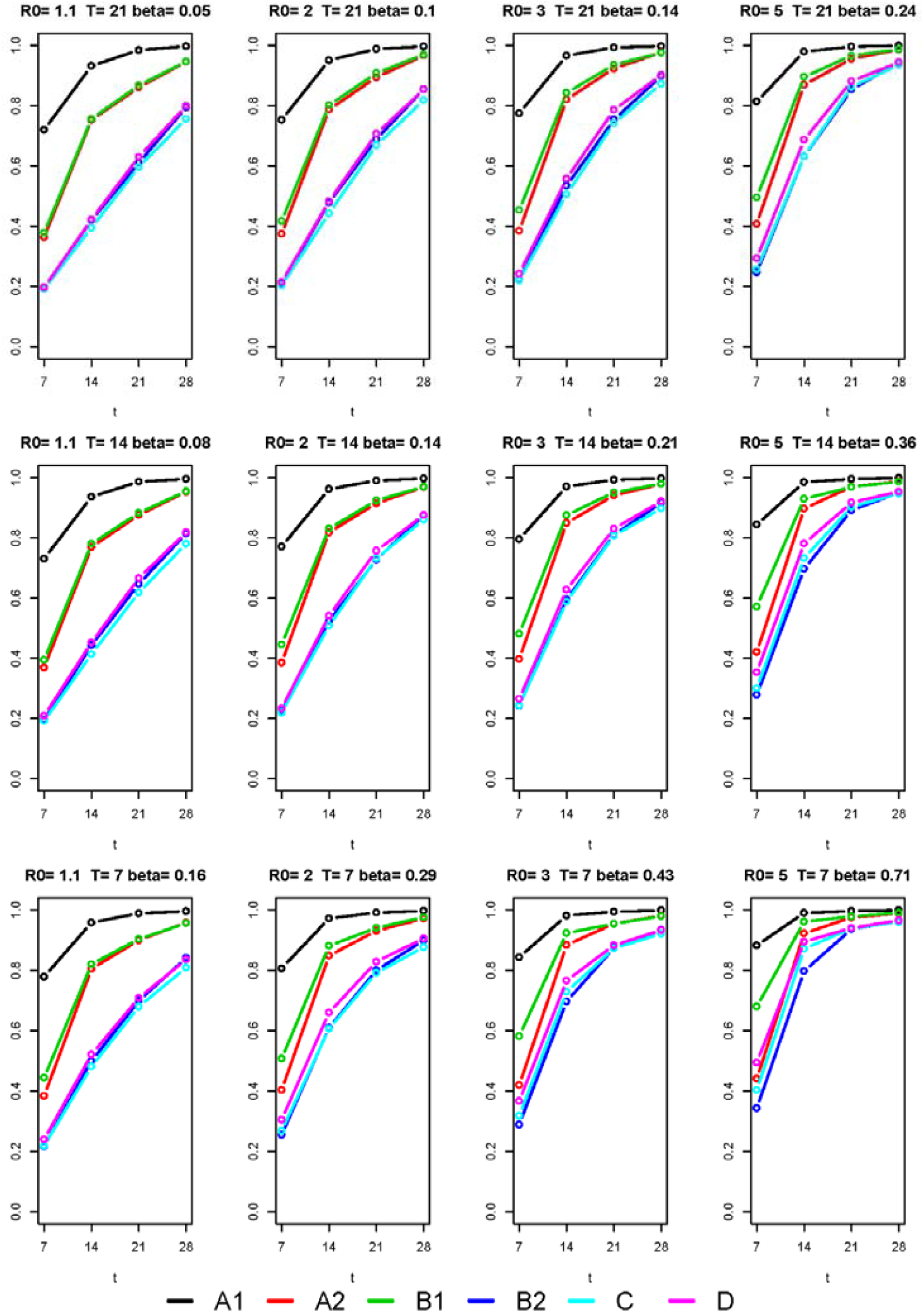
Probability of detecting the infection within t days from the beginning of the epidemic in the class. Comparison of the performance of the six screening plans (A1, A2, B1, B2, C, D), under different epidemic scenarios, assuming that individual tests have sensitivity 0.7 and maximum specificity.

The probabilities that the screening plans do not detect the infection within 4 weeks from the beginning of the epidemic are reported in Table 1. The risk of not detecting the outbreak at all decreases as the rate of infection within the class increases. For plan A1, the probability of a false negative is always negligible (lower than 0.4%). For plans A2 and B1 probabilities are very similar (from 1.0% when *β* = 0.71 to 5.5% when *β* = 0.05), as well as for plans B2, C and D (from 3.4% when *β* = 0.71 to 24.4% when *β* = 0.05).

**Table 1.** Probability of not detecting the outbreak in the class within 35 days from the beginning of the epidemic, by screening plan (A1, A2, B1, B2, C, D), under different epidemic scenarios (*R*0 and *T*), assuming that individual tests have sensitivity 0.7 and maximum specificity.

| Scenario |  |  | Screening plan |  |  |  |  |  |
| --- | --- | --- | --- | --- | --- | --- | --- | --- |
| $R_0$ | $T$ | $\beta$ | A1 | A2 | B1 | B2 | C | D |
| 1.1 | 21 | 0.05 | 0.003 | 0.055 | 0.054 | 0.206 | 0.244 | 0.201 |
| 2.0 | 21 | 0.10 | 0.004 | 0.034 | 0.031 | 0.146 | 0.182 | 0.147 |
| 3.0 | 21 | 0.14 | 0.003 | 0.024 | 0.025 | 0.102 | 0.127 | 0.099 |
| 5.0 | 21 | 0.24 | 0.001 | 0.014 | 0.013 | 0.056 | 0.065 | 0.057 |
| 1.1 | 14 | 0.08 | 0.004 | 0.047 | 0.045 | 0.185 | 0.220 | 0.181 |
| 2.0 | 14 | 0.14 | 0.003 | 0.031 | 0.030 | 0.125 | 0.139 | 0.124 |
| 3.0 | 14 | 0.21 | 0.003 | 0.021 | 0.020 | 0.083 | 0.102 | 0.079 |
| 5.0 | 14 | 0.36 | 0.001 | 0.012 | 0.014 | 0.050 | 0.053 | 0.046 |
| 1.1 | 7 | 0.16 | 0.004 | 0.042 | 0.044 | 0.158 | 0.191 | 0.164 |
| 2.0 | 7 | 0.29 | 0.003 | 0.028 | 0.024 | 0.101 | 0.123 | 0.094 |
| 3.0 | 7 | 0.43 | 0.001 | 0.019 | 0.020 | 0.066 | 0.078 | 0.066 |

Figure 3 shows, for each scenario and screening plan, the distribution of the number of lost infection-days over a time window of 4 weeks, arisen from the simulations. The colors indicate the cost of the plans in terms of individual tests. If *x* is the cost of plans B2, C and D (green), plan A2 and B1 cost 2*x* (yellow) and plan A1 costs 4*x* (red). The number of lost infection-days increases as *β* increases. After A1, B1 is the plan which assures the lowest number of lost infection-days. If we exclude scenarios of low epidemic spread, when plan B2, C and D are comparable in terms of lost infection-days, of the three cheaper plans, plan D seems the one that guarantees the lowest number of lost infection-days. Interestingly, plan D is equivalent or better than A2 in scenarios where *β* ≥ 0.36. Plan B1 leaves less undetected infection-days than plan A2.

**Figure 3.**
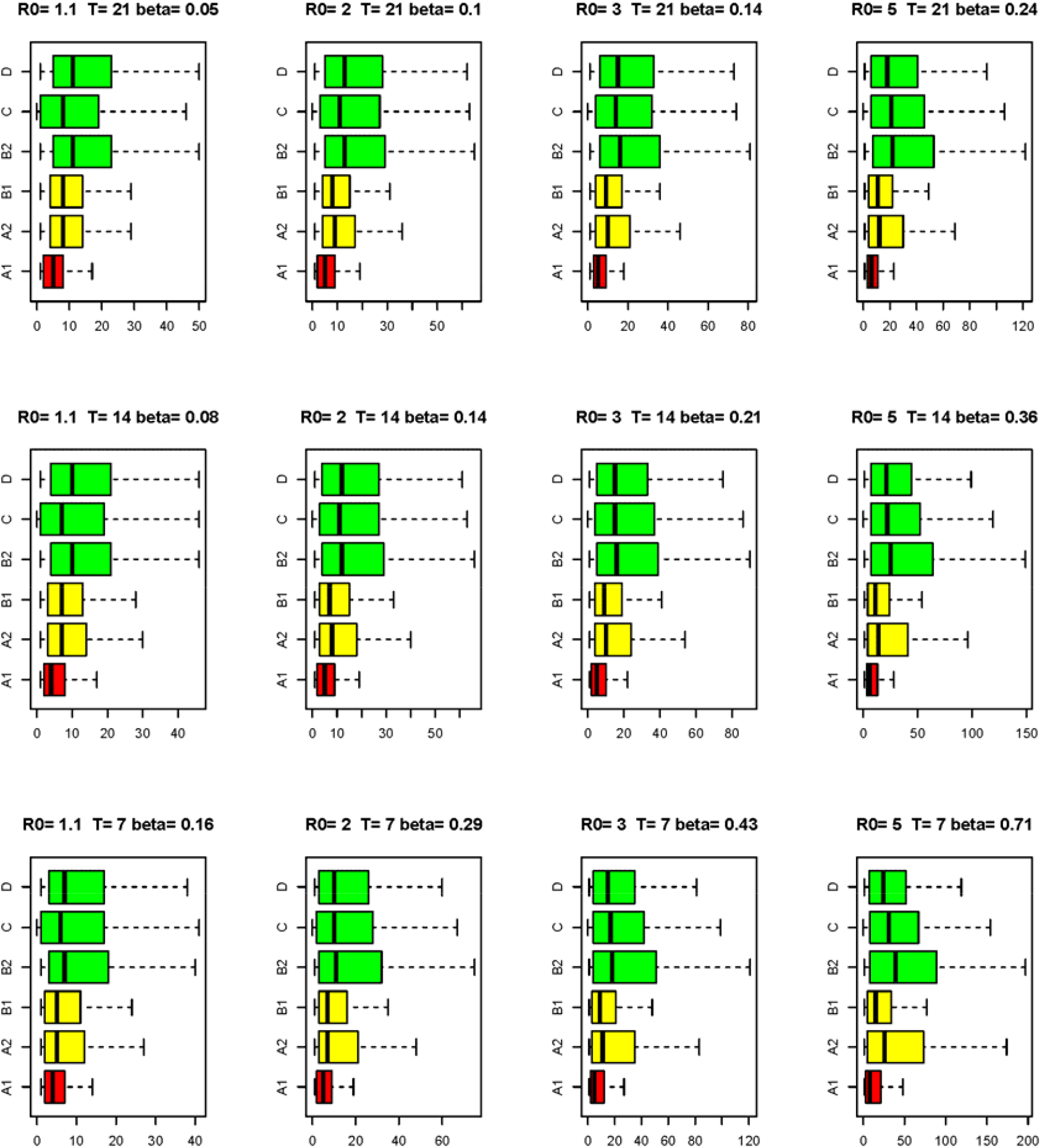
Boxplots of the lost infection-days by screening plan (A1, A2, B1, B2, C, D), under different epidemic scenarios, assuming that individual tests have sensitivity 0.7 and maximum specificity. In green the less expensive plans, in yellow the medium and in red the most expensive.

In Figure 4, focusing on plan A1 and on plans B1 and D, which are the best ones within their cost range, we compare the cumulative probabilities of a positive result when assuming *p*=0.7 and 0.9, under the scenarios characterized by the larger and the lower rates of contagion. Increasing the sensitivity of the individual test to 0.9, the performance of the screening plans increases, but their relative accuracy remains similar. This result arises also from the comparison of Tables S1 and S2 (Supplemental Material), which report averages and 90th percentiles of the number of lost infection-days for the six screening plans and the 12 epidemic scenarios, for *p*=0.7 and 0.9, respectively.

**Figure 4.**
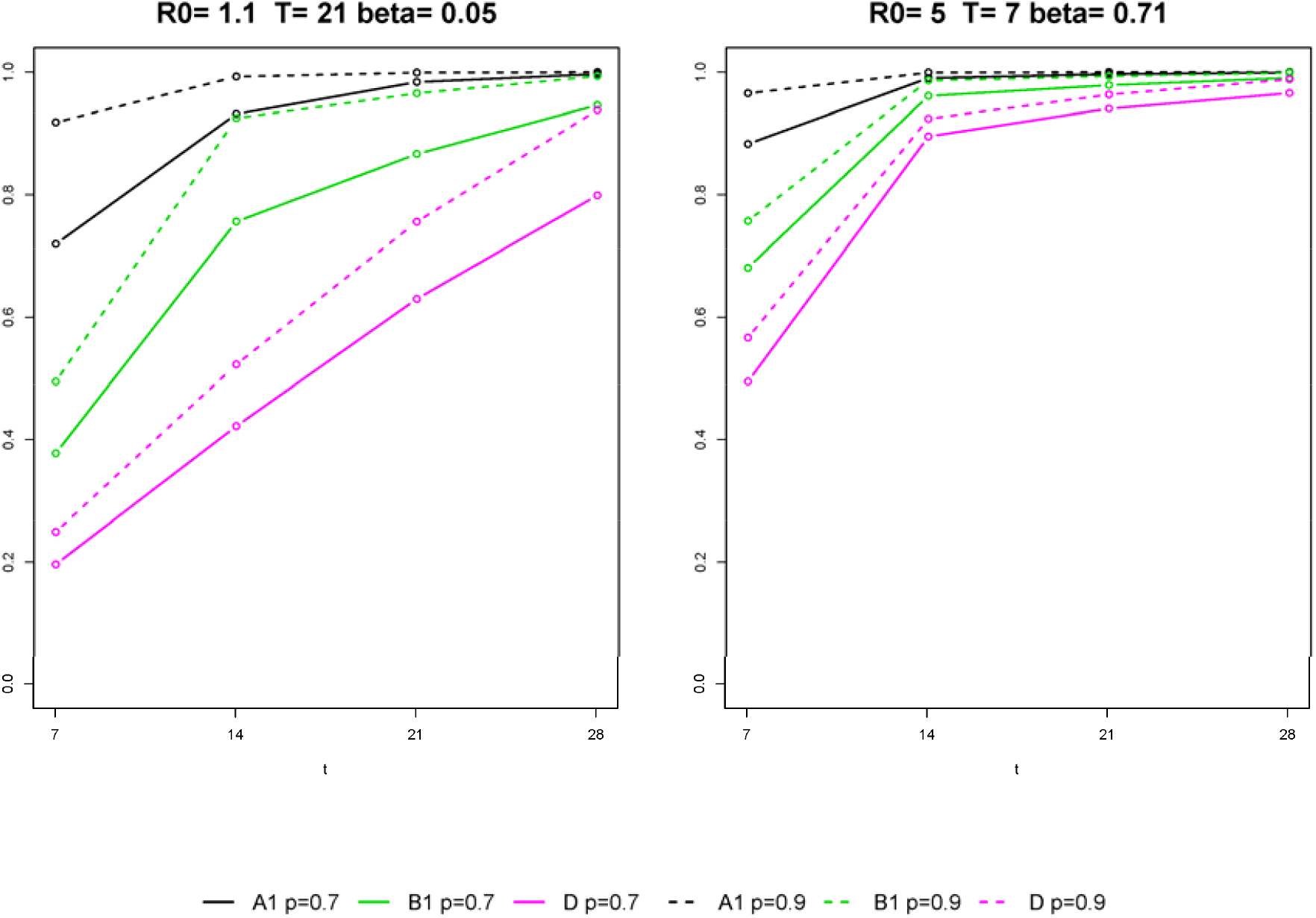
Probability of detecting the infection within *t* days from the beginning of the epidemic in the class. Comparison of the performance of the screening plans A1, B1, D, under the scenarios characterized by the lower (left panel) and the largest (right panel) rates of contagion, assuming that individual tests have sensitivity 0.7 (solid line) and 0.9 (dashed line) and maximum specificity.

## DISCUSSION

In this paper, we perform simulations to compare the performance of six screening plans based on individual tests with low sensitivity and maximum specificity in detecting infection outbreaks at the class level in schools or, more in general, at the cluster level in a population. We account for uncertainties around the epidemic dynamic in the class through a stochastic compartmental model and around the screening plan implementation through random generation of groups, when required by the plan, and random allocation of the new infections across them. Our work is not far from others which used mathematical models or simulations to investigated relevant issues during the COVID-19 emergency, such as the definition of optimal quarantine strategies [11] or optimal pool size in pooled testing [12, 13].

The compared plans have different costs and are based on recurrent evaluations on all students or on subgroups of students on rotation. Among all possible plans that perform assessments on the students at time intervals greater than one week, the best option obviously consists in testing all students at weekly intervals. This option assures the maximum sensitivity and the lowest number of infection-days left undetected. However, it is very expensive, hence the need of exploring cheaper strategies. Without claiming to be exhaustive, we consider and compare through simulations five alternatives to the best option. At an equal cost, testing half of the class on rotation every week proves to be better than testing all students every two weeks, because it allows to earlier detect the presence of infections, especially in scenarios of a high rate of contagion in the class.

If resource constraints are even tighter, less expensive plans can be considered: testing half of the class once every two weeks, testing one-third of the class every 10 days, testing one-fourth of the class every week. These plans, which have same cost, perform similarly in case of a low rate of contagion. However, in case of a high rate, testing one-fourth of the class every week seems to be the best option. This again suggest that reducing times between successive evaluations may largely balance the risk related to a lower class coverage at each assessment. Moreover, considering that strength and speed of virus transmission within each single class are not known in advance, it seems reasonable to focus on high-transmission scenarios, potentially more dangerous in terms of infection spread within and outside the class [14].

Interestingly, the plan which tests one-fourth of the students every week results to be comparable to the plan that tests all students every two weeks in terms of sensitivity and turns out to be better in terms of lost infection-days. This suggests that reducing costs by simply increasing the interval between successive assessments should not be considered a priori a good option, because much less expensive strategies based on testing subgroups on rotation could end up performing similarly.

The absolute performance of the screening plans increases with the sensitivity of the individual tests used, but their relative performance remains unchanged. It should be noticed that the sensitivity value of 0.9 roughly corresponds to the one that would be obtained by repeating a test with a sensibility of 0.7 twice on the same individual, in a procedure of double testing [6, 15].

Our simulations can be extended also to screening plans which combine rotation and sample pooling strategies, that could be less costly than those based on individual tests [16]. Each group of six students in plans C or D could be tested in pool with only one molecular test on mixed material from individual swabs. Similarly, the screening on the entire class could be done by performing only one pooled test on the whole class or by defining *k*≥1 subgroups of students, then performing *k* pooled tests. However, the sensitivity of pooled testing for various pool sizes and its relationship with viral loads distribution in the population should be considered essential inputs to assess and compare the performance of plans which involve sample pooling [17].

A final remark concerns our assumption of maximum specificity. This assumption affects the calculation of the sensitivity at the class level in a conservative sense: if a student is falsely declared positive in a class where there is at least one infected, the probability of a positive result at the class level increases. On the other hand, a suboptimal, even if high, specificity could induce a not negligible number of inappropriate quarantines, with the resulting social costs. We do not address this issue in the paper, but it is worth noting that the risk of inappropriate quarantine is lower if tests are performed on smaller groups of students. For example, if the individual test has a specificity of 0.99, the specificity on the class at each assessment is 0.99^6^=0.94 under plan C and 0.99^24^=0.79 under plan A1.

In conclusion, our simulations emphasize the importance of taking into account the epidemic context when comparing alternative screening strategies aimed at contagion prevention. Combining calculation of test accuracy with contagion dynamic models is an easy way to do this, even when the plans to be evaluated are more complex than those considered in this paper.

We show that, in the presence of clusters in the population, less costly strategies that exploit the correlation between subjects may have good performance, in particular in detecting outbreaks at high-rate of contagion, and that the time interval between successive evaluations on the same cluster is a very relevant input. The closer the assessments, the lower the number of infection-days left undetected, with a reduction of the risk that infectious subjects spread the contagion within the cluster and outside it.

In real applications, economic and social costs as well as positive and negative predictive values of the proposed plans, should be evaluated making assumptions about the prevalence of infections in the population.

## Data Availability

The paper is based on simulations.

