## Supplemental Material for "Screening plans for SARS-CoV-2 based on sampling and rotation: an example in the school setting"

| Scenario | | | Screening plan | | | | | | | | | | | |
|---|---|---|---|---|---|---|---|---|---|---|---|---|---|---|
| | | | A1 | | A2 | | B1 | | B2 | | C | | D | |
| $R_0$ | $T$ | β | mean | 90th p. | mean | 90th p. | mean | 90th p. | mean | 90th p. | mean | 90th p. | mean | 90th p. |
| 1.1 | 21 | 0.05 | 6.2 | 13 | 10.8 | 24 | 10.2 | 22 | 16.7 | 38 | 12.6 | 32 | 16.4 | 37 |
| 2.0 | 21 | 0.10 | 6.6 | 14 | 12.9 | 30 | 11.4 | 26 | 21.1 | 50 | 18.1 | 46 | 20.1 | 47 |
| 3.0 | 21 | 0.14 | 7.3 | 16 | 15.6 | 37 | 12.7 | 29 | 26.4 | 65 | 22.9 | 58 | 23.3 | 55 |
| 5.0 | 21 | 0.24 | 8.5 | 20 | 22.2 | 59 | 16.2 | 38 | 36.5 | 93 | 31.9 | 80 | 28.4 | 68 |
| 1.1 | 14 | 0.08 | 6.1 | 13 | 10.4 | 24 | 9.6 | 21 | 15.5 | 37 | 12.8 | 34 | 14.8 | 36 |
| 2.0 | 14 | 0.14 | 6.7 | 15 | 13.5 | 33 | 11.2 | 26 | 20.7 | 52 | 18.8 | 48 | 19.0 | 46 |
| 3.0 | 14 | 0.21 | 7.7 | 17 | 17.8 | 45 | 13.4 | 31 | 27.4 | 71 | 25.0 | 63 | 23.1 | 56 |
| 5.0 | 14 | 0.36 | 9.9 | 23 | 28.0 | 76 | 17.2 | 42 | 41.5 | 109 | 34.5 | 87 | 30.1 | 72 |
| 1.1 | 7 | 0.16 | 5.5 | 12 | 9.5 | 23 | 8.3 | 20 | 13.2 | 34 | 11.6 | 31 | 12.1 | 30 |
| 2.0 | 7 | 0.29 | 7.2 | 17 | 15.5 | 43 | 11.6 | 29 | 21.8 | 59 | 19.2 | 52 | 17.9 | 46 |
| 3.0 | 7 | 0.43 | 9.1 | 22 | 23.9 | 68 | 15.1 | 38 | 32.2 | 87 | 27.3 | 71 | 23.5 | 60 |
| 5.0 | 7 | 0.71 | 14.4 | 37 | 41.5 | 106 | 22.5 | 57 | 51.0 | 120 | 40.8 | 96 | 32.8 | 78 |

Table S1: Mean and 90th percentile of the number of lost infection-days by screening plan (A1, A2, B1, B2, C, D), under different epidemic scenarios ($R_0$ and $T$), assuming that individual tests have sensitivity 0.7 and maximum specificity.

| Scenario | | | Screening plan | | | | | | | | | | | |
|---|---|---|---|---|---|---|---|---|---|---|---|---|---|---|
| | | | A1 | | A2 | | B1 | | B2 | | C | | D | |
| $R_0$ | $T$ | β | mean | 90th p. | mean | 90th p. | mean | 90th p. | mean | 90th p. | mean | 90th p. | mean | 90th p. |
| 1.1 | 21 | 0.05 | 4.6 | 9 | 8.5 | 18 | 8.0 | 16 | 14.1 | 32 | 13.0 | 30 | 13.7 | 30 |
| 2.0 | 21 | 0.10 | 4.8 | 10 | 9.2 | 20 | 8.3 | 17 | 15.6 | 36 | 14.6 | 34 | 14.7 | 33 |
| 3.0 | 21 | 0.14 | 5.5 | 11 | 12.3 | 29 | 10.0 | 22 | 21.1 | 51 | 19.6 | 47 | 18.1 | 42 |
| 5.0 | 21 | 0.24 | 6.7 | 15 | 18.2 | 46 | 12.4 | 29 | 30.0 | 78 | 26.7 | 66 | 22.7 | 54 |
| 1.1 | 14 | 0.08 | 4.7 | 9 | 8.4 | 18 | 7.7 | 16 | 13.2 | 30 | 12.7 | 30 | 12.4 | 28 |
| 2.0 | 14 | 0.14 | 4.9 | 10 | 9.4 | 21 | 8.2 | 18 | 14.9 | 36 | 14.5 | 35 | 13.9 | 33 |
| 3.0 | 14 | 0.21 | 5.9 | 13 | 14.6 | 37 | 10.5 | 24 | 23.3 | 59 | 20.7 | 52 | 18.7 | 45 |
| 5.0 | 14 | 0.36 | 7.8 | 18 | 24.1 | 66 | 14.2 | 34 | 37.1 | 99 | 29.7 | 76 | 23.9 | 58 |
| 1.1 | 7 | 0.16 | 4.6 | 10 | 8.4 | 20 | 7.0 | 16 | 11.7 | 30 | 11.0 | 28 | 10.6 | 26 |
| 2.0 | 7 | 0.29 | 5.0 | 11 | 10.5 | 27 | 8.4 | 20 | 14.8 | 39 | 13.4 | 35 | 12.6 | 31 |
| 3.0 | 7 | 0.43 | 7.5 | 18 | 21.5 | 62 | 12.5 | 31 | 29.1 | 81 | 23.6 | 62 | 20.1 | 51 |
| 5.0 | 7 | 0.71 | 12.0 | 32 | 38.6 | 102 | 19.1 | 49 | 47.4 | 115 | 37.0 | 89 | 28.2 | 69 |

Table S2: Mean and 90th percentile of the number of lost infection-days by screening plan (A1, A2, B1, B2, C, D), under different epidemic scenarios ($R_0$ and $T$), assuming that individual tests have sensitivity 0.9 and maximum specificity.
